# Predicting Inpatient Glucose Levels and Insulin Dosing by Machine Learning on Electronic Health Records

**DOI:** 10.1101/2020.03.02.20029017

**Authors:** Xiran Liu, Ivana Jankovic, Jonathan H Chen

## Abstract

Poorly controlled glucose levels are associated with serious morbidity and mortality in hospitalized patients. Hospital diabetes management aims to maintain the glucose level within a desired range, primarily via insulin administration. Current inpatient glucose control relies significantly on expert knowledge, but this results in large variability and often suboptimal blood sugars in practice. We applied supervised machine learning methods to electronic health record (EHR) data to build predictive models that can inform inpatient insulin management. We found that individual blood glucose levels and insulin dosing are highly erratic and cannot be predicted precisely (MAE 28mg/dL, R^2^ 0.2). However, prescribing decisions can still be driven by the more reliable predictions of average daily glucose levels (MAE 21mg/dL, R^2^ 0.4) and whether any patient’s glucose levels will be higher than the clinically desired range in the next day (sens 0.73, spec 0.79).

## Introduction

Diabetes mellitus is a common chronic disease with serious consequences. It is a complex metabolic disorder that results from insufficient production of insulin or when the body cannot effectively use the insulin it produces.^1^ Broadly, diabetes that results from deficient insulin production is characterized as type 1 diabetes (T1DM) and diabetes that results initially from the body’s ineffective use of insulin (insulin resistance) is characterized as type 2 diabetes (T2DM). Other types of diabetes mellitus are a minority of cases and will not be addressed here. T1DM comprises 5% of diabetes and T2DM comprises 90%–95%^2^. In the United States, more than 9.4% of the population has diabetes and it was the seventh leading cause of death in 2015. The prevalence of diabetes is rising in the United States^3^.

Patients with diabetes struggle with blood sugar control. Although the pathophysiology predisposes patients with diabetes to high glucose (hyperglycemia), low glucose (hypoglycemia) can also occur due to inappropriate dosage of anti-diabetic medications. Both hyperglycemia and hypoglycemia can have a life-threatening impact. Acutely, severe hypoglycemia can lead to seizures, loss of consciousness, or death. In the long term, diabetes leads to complications of blindness, kidney failure, lower limb amputation, increased risk of cardiovascular disease, and other conditions that significantly impact health and quality of life^1^.

According to the standards used by American Diabetes Association (ADA), the International Expert Committee (IEC), and the World Health Organization, an A1c (glycated hemoglobin test) percentage ≥6.5% (≥48 mmol/mol), fasting plasma glucose ≥126 mg/dL (milligrams per deciliter), and glucose ≥200 mg/dL measured 2 hours after an oral glucose load are diagnostic for diabetes^2^. In hospitalized patients, hypoglycemia (low blood glucose) is generally considered a glucose ≤70 mg/dL (3.9 mmol/L)^4^.

Hospital goals for the patient with diabetes focus on the management of blood glucose levels to prevent glycemic excursions.^5^ Although there are many medications used for outpatient blood sugar control, especially in T2DM, insulin alone is the standard treatment for inpatient diabetes management^6^.

Insulin is available in various forms, characterized by rapidity of onset after injection, peak in functioning, and duration of action. Rapid- and short-acting insulin have a rapid onset and peak in a few hours, whereas intermediate- and long-acting insulin can work for as long as approximately 48 hours^7^.

The use of a basal-bolus insulin regimen for the management of hyperglycemia is recommended by many studies^8^. Basal insulin, typically a long-acting insulin given once or twice a day, is given to keep blood glucose at consistent levels during periods of fasting and to cover non-prandial insulin needs. Short-acting bolus insulin is given to control glucose loads following a meal. “Sliding scale” insulin (SSI), in which regular insulin or a rapid-acting insulin analogue is dosed by blood glucose increments (e.g. one unit of insulin for every 50mg/dL that the glucose is >150mg/dL) can be given in addition to basal-bolus regimens or without any other scheduled insulin^9^. Some form of SSI therapy has been around since the 1930s and is popular in hospitals^10^. However, because it treats hyperglycemia after it had already occurred, SSI alone is a reactive strategy, and is not effective at glucose control^8,11^, especially when compared to a basal-bolus regimen in hospitalized diabetic patients^8,12^. Nonetheless, suboptimal insulin regimens remain common in medical inpatients with diabetes mellitus^13^.

Electronic health record (EHR) tools for diabetes management are believed to have the ability to improve diabetes patient outcomes through enhanced education, patient support, and reduced clinical inertia on the part of the healthcare provider^14,15^. There are also studies on probabilistic models and machine learning approaches for blood glucose prediction and diabetes management^16–18^. However, prior work either analyzes data only from a single patient^17^, or uses physiological model with experimental parameters that need manual work when registering data and tuning model to the individual patient^16,18^. The level of manual work required and the detailed analysis of individual patients are inefficient in practice. In this study, we work with de-individualized prediction models that do not require knowledge of physiological parameters or any manual work other than the collection of EHR data, thus they are more generalizable and feasible in practice.

Hospital glucose control aims to maintain the glucose level within 140-180 mg/dL^19^, principally via insulin dosing. Inpatient glucose management relies significantly on expert knowledge and manual review of inpatient glucose. Although some experienced doctors order basal-bolus insulin regimens effectively, glucose control dependent on humans is globally variable and thus suboptimal. There is therefore the potential to improve inpatient care quality through the use of learning methods to augment decision support in EHRs.

## Objective

The objective of this project is to determine how well inpatient glucose levels and the amount of insulin ordered can be predicted through supervised machine learning methods applied to electronic health records.

## Data Processing

The dataset used contains EHR data from STARR (STAnford Research Repository) Datalake. For the purpose of this project, only records after January 1st, 2014 are considered. The study was approved by the Stanford Institutional Review Board.

Patients with the chance to be diagnosed as having hyperglycemia (or hypoglycemia) were first selected from the entire datalake using the criteria of having a glucose level measured ≥ 200 mg/dL or ≤ 70 mg/dL or having A1c percentage ≥ 6.5%. Approximately 42,700 patients were selected. This project focused on patients receiving subcutaneous insulin. Patients were excluded if important information, including patient weight, was missing, or if there are fewer than five glucose measures within 72 hours. To avoid unnecessary complexity, we also excluded patients on hemodialysis. After filtering on the above criteria, 3,461 patients remained in the dataset. Imputation methods were not considered because inferring information such as weight from other measures, e.g., glucose levels, may cause unwanted dependency in prediction, and inferring glucose measures is the goal of our prediction model.

Prior reports on diabetes have shown that the number of diagnosed and undiagnosed patients with diabetes, as well as morbidity attributed to diabetes, varies for each age-sex group^1,3^. These studies suggest that that age and sex may provide useful information, so both date of birth and gender, provided in the demographic dataset of the datalake, were extracted.

We used several lab results from EHRs, including glucose values by (gluco)meter, A1c percentage, and creatinine. Creatinine, a measure of kidney function, can be helpful for indicating impaired insulin clearance. Elevated creatinine level signifies impaired kidney function.

Orders of insulin and glucocorticoids were also included. In this project, insulin was classified into two categories: short- and long-acting, based on its speed and length of functioning. The short- (and rapid-) acting category contains regular insulin (to clarify: “regular” is a type of insulin), Lispro, and Aspart. The long- (and intermediate-) acting category contains NPH, Detemir, and Glargine^5^. At each timestamp, the total short-acting dose within past half hour, 1 hour, 3 hours, and 6 hours was calculated based on the record of medication orders, and similarly the total long-acting dose within the past 6 hours, 12 hours and 24 hours. Glucocorticoids are widely-used anti-inflammatory and immunosuppressive drugs. They are also the drug group most often associated with the onset of hyperglycemia^20,21^. Four major systemic glucocorticoids and their equivalent doses are hydrocortisone (20 mg), prednisolone (5 mg), methylprednisolone (4 mg), and dexamethasone (0.75 mg) ^22^. The amount of glucocorticoid ordered within 24 hours and three days were both calculated, with the amount of each type of glucocorticoid transform converted to standard units based on the dose equivalence then summed together. We also considered other potentially relevant information. Because glucose comes from food, diet of patients would affect glucose levels. It is challenging to capture “eating” behaviors from the EHR, however we did consider information on diet status, i.e., whether patients were NPO (nothing through the mouth). The relative time in a day of predicted glucose was also incorporated to capture diurnal patterns of glucose and the insulin order set, which is by default three times a day before meals and at bedtime.

With information containing the characteristics above for each patient at each timestamp, the feature space consisted of 20 features, summarized in Table 1. There were 175,934 data points after filtering (from 3,461 patients), each corresponding to a timestamp. The three values we were interested in predicting were also extracted: the current glucose level, the average glucose level for next 24 hours, and the total amount of insulin ordered in next 24 hours from the current timestamp. The percentages of blood glucose measures of the patients in the records that are high, low, and normal are shown in the pie chart in Figure 1. Although the threshold for high glucose level by which we chose to filter the data, ≥ 200 mg/dL, is more liberal than the inpatient maximum target of 180 mg/dL, 25% of the blood glucose levels were above our threshold. At approximately 1.7% of the timepoints, patients were hypoglycemic. From the plot of average glucose levels against insulin ordered in next 24 hours from each timestamp shown in Figure 1, we observed that the actual insulin ordered in the hospital for a given glucose varied widely, suggesting wide practice variation. This large variance in insulin ordered results from the difficulty in physicians’ prediction of future blood glucose, and, by extension, of appropriate insulin doses.

**Table 1.**
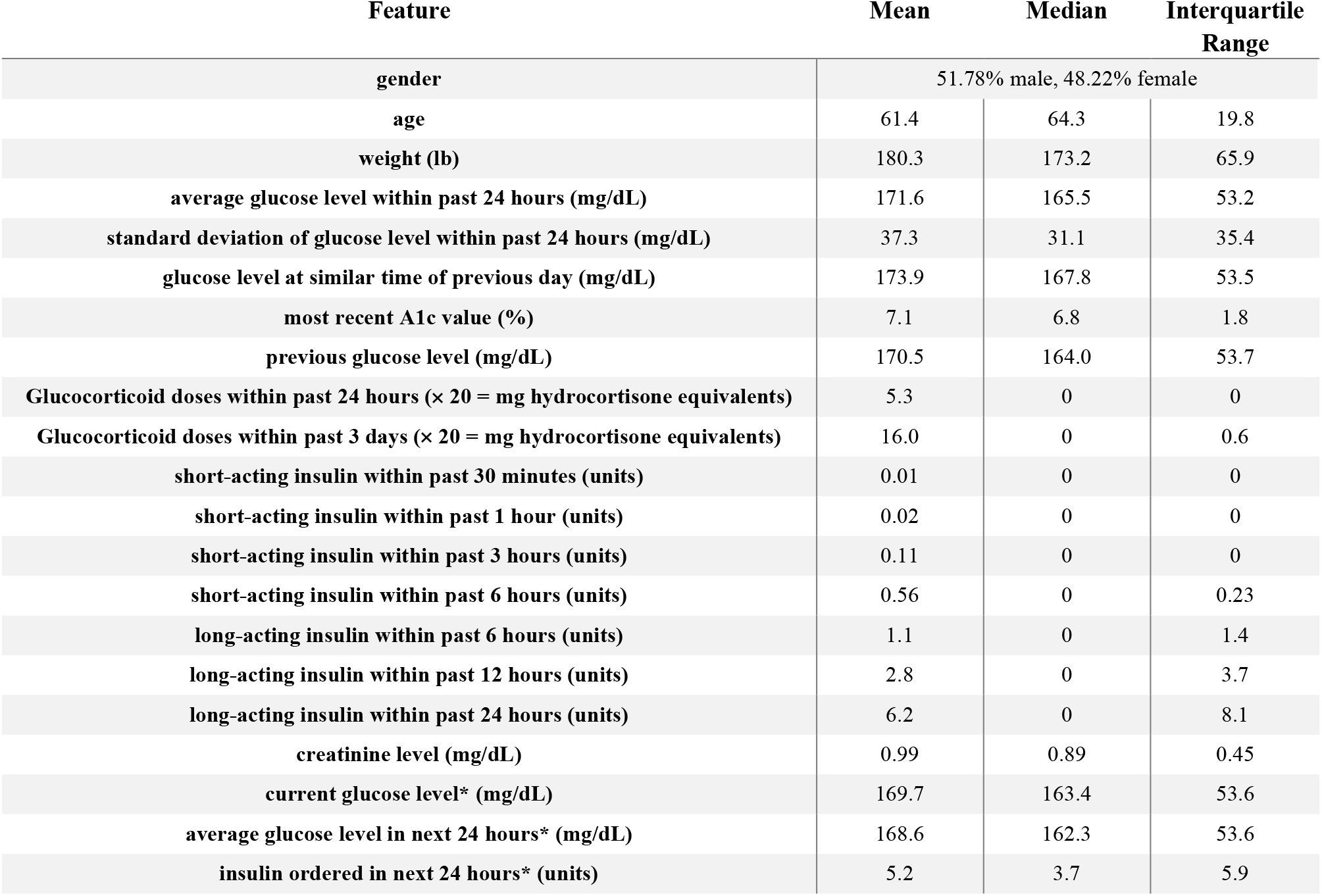
Statistics of extracted features: mean, median and interquartile range (difference between 75th and 25th percentiles); asterisk symbol denotes values to be predicted

| Feature | Mean | Median | Interquartile Range |
| --- | --- | --- | --- |
| gender | 51.78% male, 48.22% female |  |  |
| age | 61.4 | 64.3 | 19.8 |
| weight (lb) | 180.3 | 173.2 | 65.9 |
| average glucose level within past 24 hours (mg/dL) | 171.6 | 165.5 | 53.2 |
| standard deviation of glucose level within past 24 hours (mg/dL) | 37.3 | 31.1 | 35.4 |
| glucose level at similar time of previous day (mg/dL) | 173.9 | 167.8 | 53.5 |
| most recent A1c value (%) | 7.1 | 6.8 | 1.8 |
| previous glucose level (mg/dL) | 170.5 | 164.0 | 53.7 |
| Glucocorticoid doses within past 24 hours ($\times 20 =$ mg hydrocortisone equivalents) | 5.3 | 0 | 0 |
| Glucocorticoid doses within past 3 days ($\times 20 =$ mg hydrocortisone equivalents) | 16.0 | 0 | 0.6 |
| short-acting insulin within past 30 minutes (units) | 0.01 | 0 | 0 |
| short-acting insulin within past 1 hour (units) | 0.02 | 0 | 0 |
| short-acting insulin within past 3 hours (units) | 0.11 | 0 | 0 |
| short-acting insulin within past 6 hours (units) | 0.56 | 0 | 0.23 |
| long-acting insulin within past 6 hours (units) | 1.1 | 0 | 1.4 |
| long-acting insulin within past 12 hours (units) | 2.8 | 0 | 3.7 |
| long-acting insulin within past 24 hours (units) | 6.2 | 0 | 8.1 |
| creatinine level (mg/dL) | 0.99 | 0.89 | 0.45 |
| current glucose level* (mg/dL) | 169.7 | 163.4 | 53.6 |
| average glucose level in next 24 hours* (mg/dL) | 168.6 | 162.3 | 53.6 |
| insulin ordered in next 24 hours* (units) | 5.2 | 3.7 | 5.9 |

**Figure 1.**
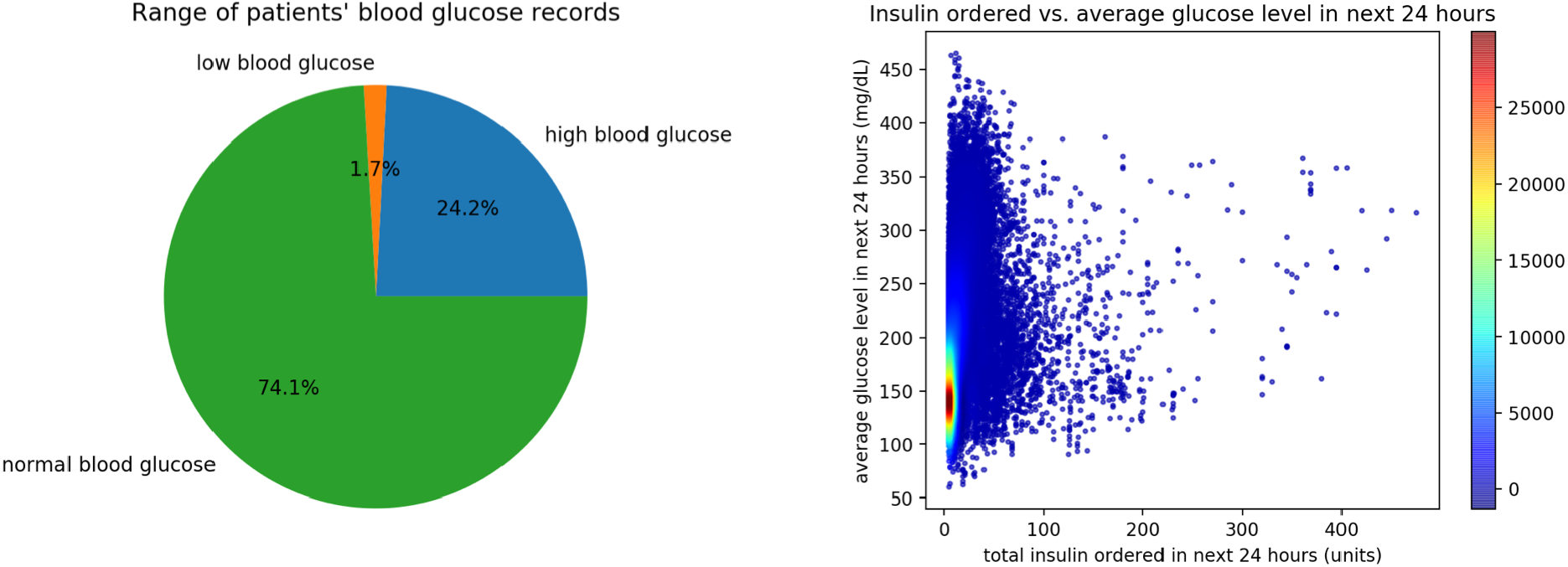
Left: Distribution of glucose measurements characterized as low (≤ 70 mg/dL), normal (71-199 mg/dL), or high (≥ 200 mg/dL). Right: Average glucose in 24 hours (mg/dL) plotted against the total insulin doses ordered in those 24 hours; unit of heatmap is number of individual measures. Both illustrate the substantial fraction and distribution of observed glucose levels outside of desired ranges.

## Methods

The problems of predicting inpatient glucose and the amount of insulin ordered in the next 24 hours based on data provided in EHRs can both be modeled as regression problems in supervised machine learning. This section describes methods used in designing and implementing procedures to address the problem.

The goal of supervised machine learning is to learn a mapping from inputs x to outputs y given a labeled set of input-output pairs (i.e., training set) {(x_i_, y_i_)_i=1,…,n_}. Each training input (data point) contains x_i_, a vector of values called the feature vector. It also contains an associated label or value y_i_ that indicates the attribute of the data point. When y_i_ is continuously-valued, the problem is known as regression. When y_i_ is categorical, the problem is known as classification. Regression models a target prediction value (output) based on given set of variables (inputs). It is a supervised learning technique commonly used in forecasting and studying relationships between variables. There are many types of regression algorithms. This project mainly focuses on two commonly-used regression algorithms: the support vector regression (SVR) algorithm and the tree-based random forest (RF) regression algorithm. Other commonly used methods including Elastic Net, Lasso, decision tree and Bayesian ridge regression were also tested, however they yielded comparable performances to SVR and RF so we focused on the former methods here.

In the problem of predicting inpatient glucose levels, the inputs were features constructed from selected data in the EHR up to the time at which we want to make the prediction. Two outputs were then considered. The first output was the glucose level at that time, and the second output was the average value of glucose levels in the next 24 hours. In the problem of predicting amount of insulin ordered, the inputs were similar to those for the prediction of inpatient glucose levels, and the output was the amount of insulin ordered in the next 24 hours.

The basic construction of features followed the steps described in last Section. Pearson correlation showed that the features were all correlated with the target output. Pearson’s correlation coefficient, which indicates the extent to which two variables are linearly related, is the covariance of the two variables divided by the product of their standard deviations. It has a value between −1 and 1. The formula for the sample correlation coefficient used in this step is 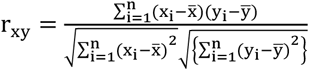 for feature x and output y. The correlations between the target output and some of the features were very weak, so we considered feature selection, which keeps only features that are relatively highly-correlated with the output to reduce the run time of learning algorithms.

For prediction of current glucose levels, we performed feature selection based on univariate linear regression test to select best features. The top five features selected out by this process were: average glucose level in past 24 hours, glucose level at the similar time of previous day, A1c percent, variance in glucose level in past 24 hours, and short-acting insulin dosing in past 3 hours. The top ten features selected in the same way are summarized in Table 2. These features are expected to have a relatively large influence on the current glucose level of a patient. In performing regression using only the top five selected features versus all the features, the experimental results showed that performing feature selection reduced the run time of algorithms, but also slightly lessened the quality of prediction.

**Table 2.**
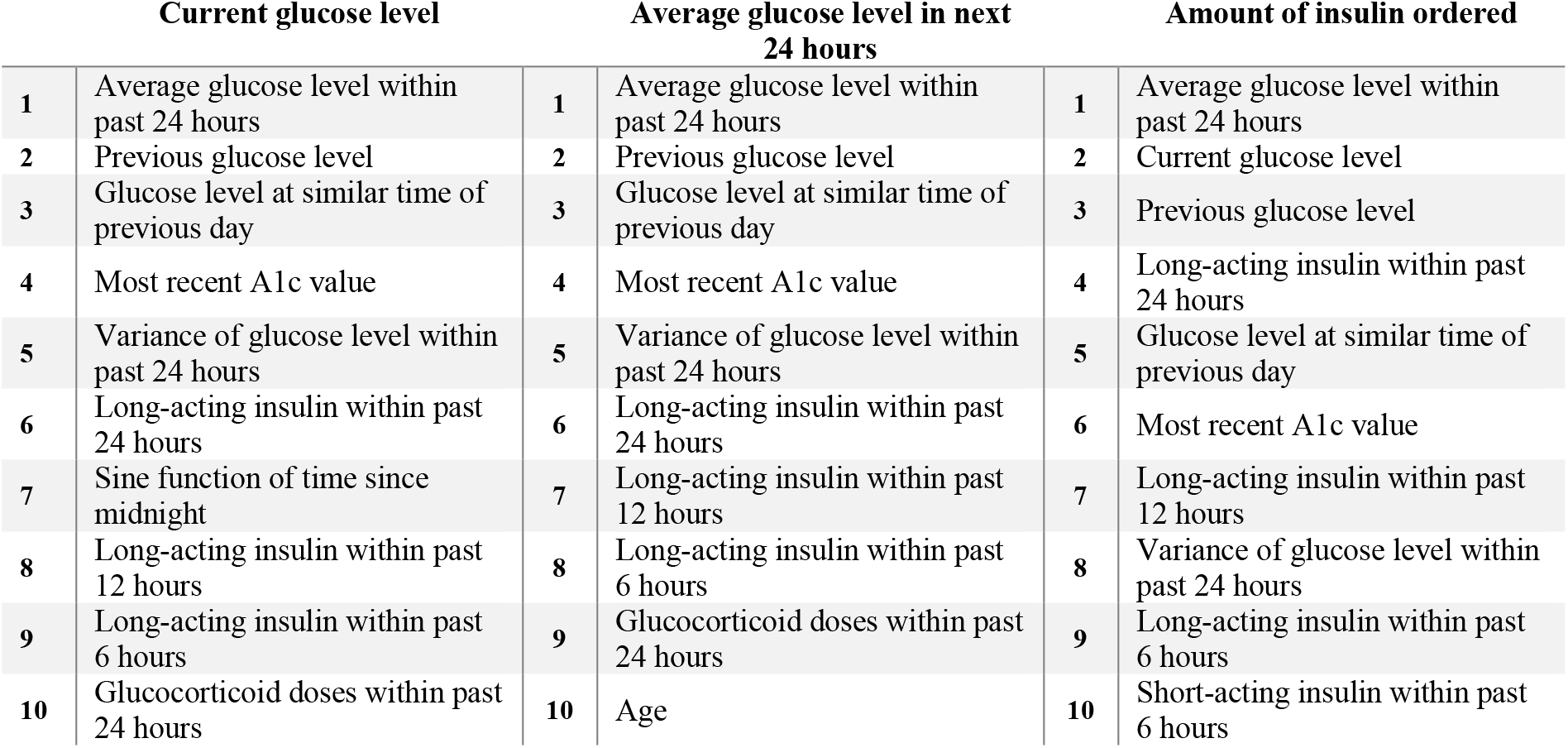
Top ten features selected for each output based on univariate statistical tests

The top ten features selected for daily average glucose level, average glucose in the next 24 hours, and insulin dosing amount are summarized in Table 2. The top five selected features for amount of insulin that clinicians ordered in the next 24 hours were: long-acting insulin doses in the past 24 hours, the past 12 hours, the past 6 hours; average glucose level in past 24 hours; and the current glucose level. Similarly, the results show that the performance was higher for models taking in all features, while those using only a subset of features had shorter run time. For the purpose of this project, because running time of the algorithms was not a big issue, we kept all features to incorporate as comprehensive information in the model as possible for a better performance.

In machine learning, data are usually split into a training dataset and a testing dataset. In this project, data were split into training and testing sets according to a ratio of 7:3 in number of patients instead of data points, which corresponded to the different number of timestamps at which glucose level was measured or insulin ordered was calculated. Each “patient” contained different numbers of data points, so the resulting size of training and testing set was not exactly 7:3, but this splitting method worked well for the purpose of training a model and testing its performance. Using data from 1000 patients, the training set contained around 33,000 data points and the testing set around 16,000 data points. The parameters for each model were tuned on a separate validation dataset. Validation provides an unbiased evaluation of a model’s fit on the training dataset while tuning the model’s parameters.

We use two evaluation metrics to measure the prediction performance of regression. The first metric is the most straightforward: the mean absolute error (MAE). Denoting the true output values (glucose levels or amount of insulin) as y_i_’s and predicted ones as 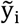’s, with each i = 1,2, …, n as indices of n data points, MAE is defined as 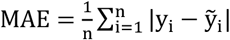. The second evaluation metric is the R^2^ score (coefficient of determination). It is a scale-free metric that measures the proportion of the variance in the dependent variable that is predictable from the independent variables. It equals one minus the ratio between the sum of squares of errors and the total sum of squares, which is proportional to the variance of the data. R^2^ score provides an indication of good fit and a measure of how well unseen observations are likely to be predicted by the model through the proportion of explained variance. It is defined as 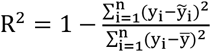, where 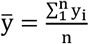 is the mean of true outputs. The best possible score for R^2^ is 1, when outputs are perfectly predicted and the errors between prediction and true values are zero for all data points. R^2^ can be negative since the model can be arbitrarily worse, as when the mean of the data provides a better fit to the outcomes than do the fitted function values (the predicted outputs). R^2^ = 0 when a model constantly predicts the mean of the data, disregarding the input features.

The problem of determining whether inpatient glucose level is high or not can be modeled as a binary classification problem in supervised machine learning. Classification problems with labels belonging to {0,1} belong to binary classification. In this scenario, the two classes are “high glucose level” and “non-high glucose level”, corresponding to glucose level at 200 mg/dL and above and glucose level below 200 mg/dL. These values are chosen based on diagnosis standard, as introduced in Section 1.1. Low glucose level, although clinically important, was not considered separately here because the number of data points corresponding to low glucose was small (only 1.7). A binary classification problem aims at classifying unknown observations into given sets of categories, on the basis of knowing a set of data containing observations with known category. Similar to regression problems, two commonly used classification algorithms are used: support vector machine (SVM) and random forest classification (RFC). The data were unbalanced because these categories contain different numbers of data points-the number of non-high glucose levels was about three times as large as that of high ones. To handle this imbalance, weighting was used in the algorithm. Data points in the smaller category received higher weights during training^23^.

The evaluation metrics used for the performance of classification are accuracy, sensitivity, and specificity. Accuracy is the number of correct predictions over the total number of predictions. Sensitivity (also called true positive rate) is the proportion of actual positives that are correctly identified, which equals the true positive predictions over the sum of true positive and false negative predictions. Here, true positives correspond to the number of positively-labeled data points, namely, high glucose levels, that are predicted correctly. False negatives are the true high glucose levels that are predicted to be non-high. Specificity, also called true negative rate, is the proportion of actual negatives that are correctly identified, which equals the true negative predictions over the sum of true negative and false positive predictions. Denoting the true output labels (whether high glucose level or not) as y_i_s, where y_i_ = 1 for high glucose level and y_i_ = 0 for non-high, and predicted ones as 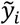s, with each i = 1,2, …, n as indices of n data points, these metrics are defined as 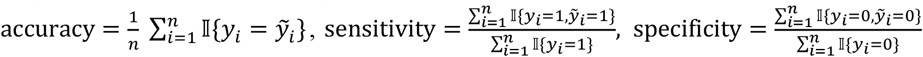, where 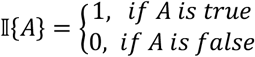 is the indicator function. Higher values of these measures indicate better classification performances.

## Results

The results for predicting current glucose level and those of predicting average glucose level in next 24 hours are summarized in Table 3. Scatter plots of mean prediction values for glucose level using SVR model against the actual values are shown in Figures 2 and 3, and the plot for prediction of insulin ordered is displayed in Figure 4. Performances of both regression algorithms are summarized in Table 3. As an example, a series of consecutive glucose levels predicted for an individual patient over a period of time is shown in Figure 5. The red points are predicted glucose levels and the blue points are true inpatient glucose levels. The top panel shows the prediction of the RF algorithm and the bottom panel shows that of the SVR algorithm. The results for classification of glucose level are summarized in Table 4. Again, the two algorithms yielded similar performances in terms of accuracy, sensitivity, and specificity.

**Table 3.**
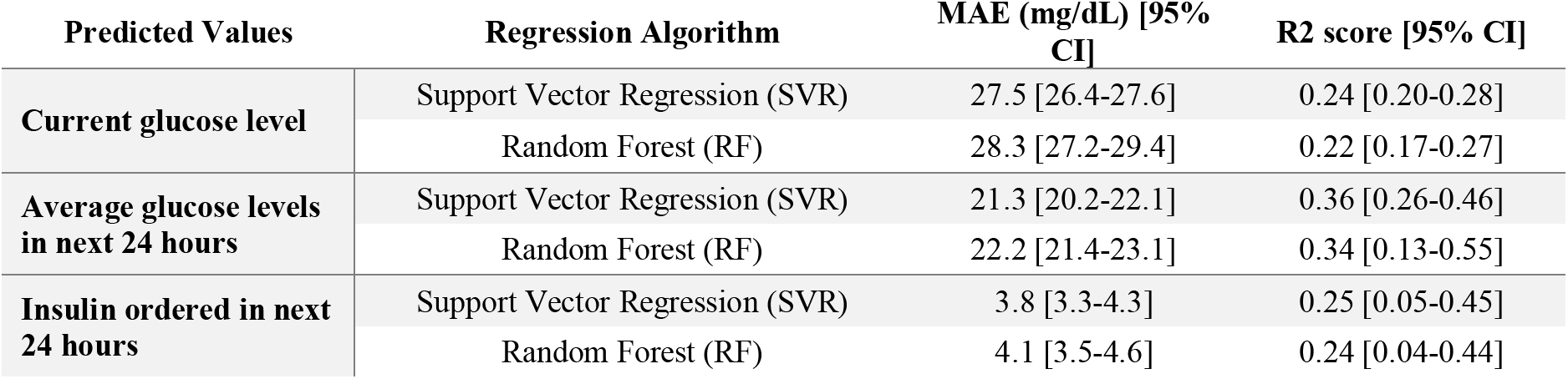
Regression performance of predicting glucose levels, average glucose levels in next 24 hours and insulin ordered in next 24 hours, assessed by mean absolute error (MAE) and R^2^ score (coefficient of determination) with 95% confidence interval (CI)

**Table 4.** Classification performance of determining whether the next glucose is high (≥ 200 mg/dL), assessed by accuracy, sensitivity (true positive rate), and specificity (true negative rate) with 95% confidence interval (CI)

| Classification Algorithm | Accuracy [95% CI] | Sensitivity [95% CI] | Specificity [95% CI] |
| --- | --- | --- | --- |
| **Support Vector Machine (SVM)** | 0.78 [0.76-0.79] | 0.73 [0.70-0.75] | 0.79 [0.77-0.81] |
| **Random Forest Classifier (RFC)** | 0.75 [0.74-0.77] | 0.79 [0.77-0.81] | 0.74 [0.72-0.76] |

**Figure 2.**
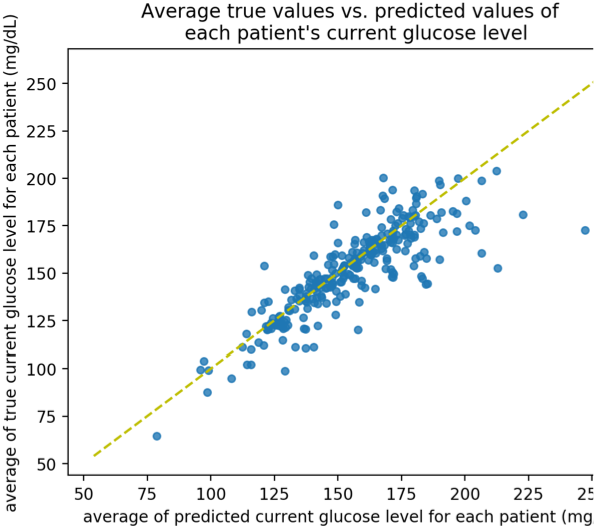
Scatter plot of SVR performance: true vs. predicted average current glucose level for each patient.

**Figure 3.**
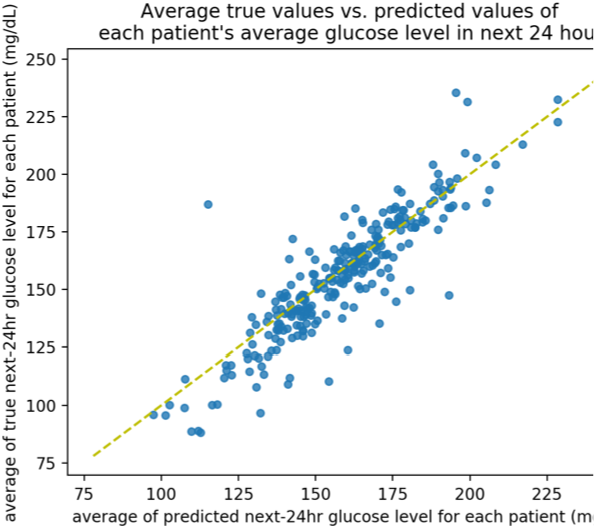
Scatter plot of SVR performance: true vs. predicted value of mean glucose level in next 24 hours for each patient

**Figure 4.**
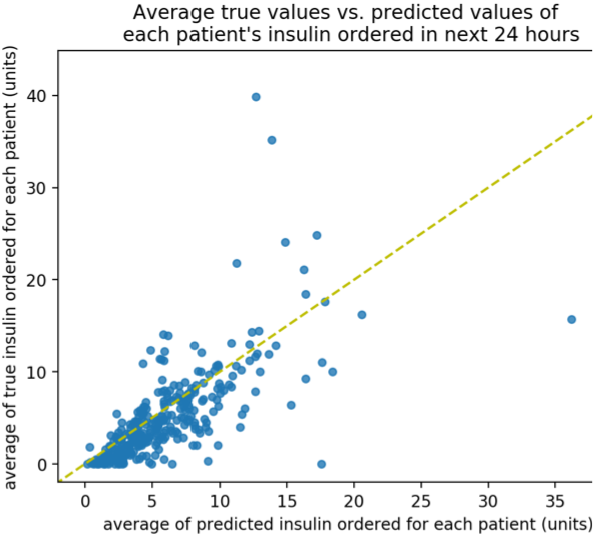
Scatter plot of true vs. predicted average of insulin ordered in next 24 hours for each patient

**Figure 5.**
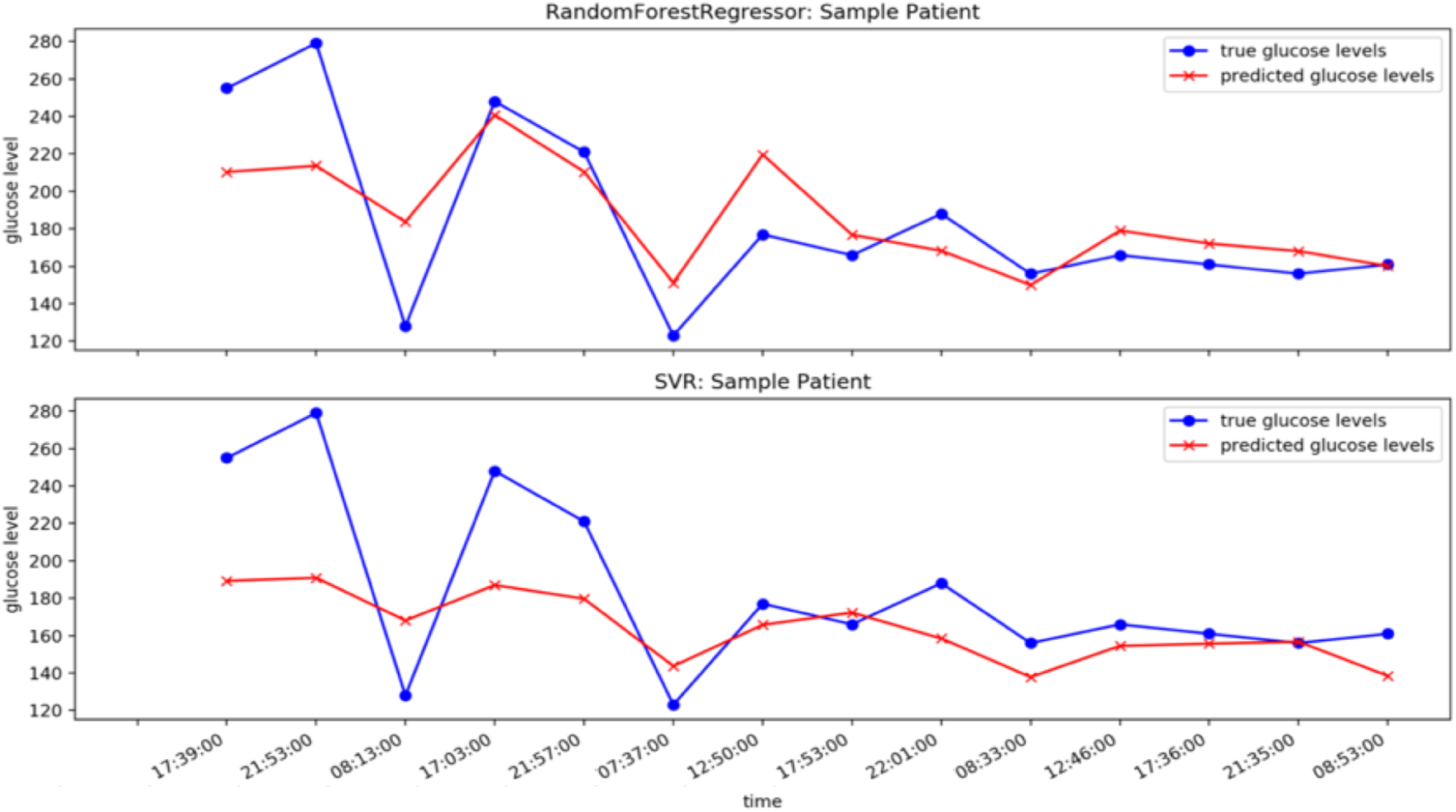
Predicted and true glucose levels over time of an individual patient using two different regression models. Red points are predicted glucose levels and blue points are true inpatient glucose levels. The top panel shows the prediction of the RF algorithm and the bottom panel shows that of the SVR algorithm. The up-and-down lines illustrate the highly variable nature of individual glucose measurements.

The mean absolute error of predicted current glucose levels was 27.5 mg/dL with a 95% confidence interval (CI) of +/−1.1 mg/dL for using SVR model and 28.3 mg/dL for using RF model with a 95% CI of +/−1.1 mg/dL, where the range of desired glucose level was 70-200 mg/dL and the mean current glucose level in the testing dataset was 163.2 mg/dL. The R^2^ score (coefficient of determination) of prediction was 0.24 for using SVR model and 0.22 for using RF model. Although R^2^ score does not indicate whether a regression model is adequate, it serves as a useful dimensionless measure of how much of variation is explained by the prediction model out of the total variation of data and compensates our measure of accuracy using mean absolute error nicely. There were no significant differences between two regression algorithms in terms of MAE and R^2^ score. These R^2^ scores imply that around 22% and 24% of the variability of the dependent variable, namely the current glucose level, had been accounted for by the corresponding model.

In learning problems, performance of a model relies not only on the algorithms itself but also on the characteristics of dataset. For example, the high variance of individual glucose levels, as indicated by the large interquartile range in Table 1, makes it challenging to make precise predictions of individual values.

Smoothing refers to capturing important patterns in the data while leaving out noise or other fine-scale structures and is naturally involved in regression. Smoothing in regression tends to reduce the variance, so the predicted values are less erratic comparing to the true values in the testing dataset. The standard deviation of current glucose level in the testing dataset was 59.5 mg/dL. This high variance of the dataset, together with the nature of regression algorithms, partially accounts for loss of predictability in this problem.

For predicted average glucose level in next 24 hours, the mean absolute error was 21.3 mg/dL with a 95% CI of +/−0.9 mg/dL for using SVR model and 22.2 mg/dL for using RF model with a 95% CI of +/−0.8 mg/dL, where the mean average glucose level in next 24 hours in the testing dataset was 162.6 mg/dL. The standard deviation of average glucose level in next 24 hours was 45.8 mg/dL. The R^2^ score (coefficient of determination) of prediction was 0.36 for using SVR model and 0.34 for using RF model, which implies that around 36% and 34% of the variability of the dependent variable, namely the average glucose level in next 24 hours, had been accounted for by the corresponding model. These values were higher than those of the predicted current glucose level, which indicates higher accuracy in the prediction. Compared to taking a single value of the glucose at the current timestamp, taking the average value of glucose measured in next 24 hours reduced the variance and thus improved the ability of capturing the correct patterns in data.

The mean absolute error of predicted amount of insulin ordered in next 24 hours was 3.8 units of insulin with a 95% confidence interval (CI) of +/−0.5 unit for using SVR model and 4.1 units of insulin for using RF model with a 95% CI of +/−0.5 unit, where the mean amount of insulin ordered in testing dataset was 5.8 units with a standard deviation of 9.4 unit. Again, there were no significant differences between the two regression algorithms in terms of MAE and R^2^ score, and the variance of the testing dataset was large.

## Discussion

The results of feature selection shown in Table 2 show that past glucose levels, A1c values, and insulin dosing are among the most relevant features to predicting glucose level and insulin, which is as expected from the clinical perspective. Although weight-based dosing is recommended by guidelines for insulin treatments, weight was surprisingly not one of the main features for amount of insulin ordered, which suggests that the guidelines may not always be followed in diabetes management and further demonstrates the suboptimality of inpatient glucose control.

The inpatient glucose level at a single timestamp was predicted through supervised machine learning methods applied to EHRs with a mean absolute error of approximately 28 mg/dL. Prediction of the average of inpatient glucose levels in 24 hours and classification of the current glucose level as being high vs. normal attained better performance in terms of accuracy. The average inpatient glucose level was predicted with a mean absolute error of approximately 22 mg/dL and about 35% of the variability accounted for by the corresponding model. High glucose levels can be distinguished from not-high ones with an accuracy of 77%. The erratic nature of the data makes the prediction of glucose level challenging. Nonetheless, the predictive models can still capture some patterns in the data and yield the correct general trend of these values. Exact prediction of individual glucose level is desired but not necessarily needed for clinical decision making. For example, continuous glucose monitors have errors of ∼9-20% in measuring current blood glucose, upon which value patients make insulin dosing decisions^24,25^. This mean absolute relative difference for continuous glucose monitors is not dissimilar from our MAE, suggesting our predictor could offer clinical guidance. Prediction of average glucose level and classification of the range of glucose level are more feasible, and the information provided by these predictions is still useful guidance for clinicians.

The mean absolute error of predicted amount of insulin ordered by clinicians in next 24 hours was around 3.8 units of insulin with about 25% of variability accounted for by the model. As shown in Figure 1, the insulin ordered by doctors in practice may fail to bring the blood glucose to a desired level. Inpatient glucose control relies significantly on expert knowledge and manual review of inpatient glucose, and these personalized experiences and decision-making processes can be hard for automated models to predict. The predicted values did not mimic the amount of insulin ordered by doctors very accurately, but these depersonalized predictions may have some implication in the potential amount of insulin needed.

The project has several limitations. We only worked with structured, de-identified EHR data, and with patients that have at least a few initial, consecutive records in the dataset. The patients are required to have all the important information, including date of birth, gender, weight, etc. Patients with missing data are not considered in this project, but missing data may happen in practice and can make the task of data processing and feature embedding more complicated. It should also be noted that our dataset focuses on patients with regular glucose level checks. They were potentially more likely to have poorly-controlled blood sugars because the need for monitoring was recognized, and this potential bias may make our findings less generalizable to a broad population. In the problem of predicting insulin ordered, given the information in EHRs, the model is only predicting the insulin prescribed by clinicians, but not necessarily the actual amount of insulin needed for maintaining the patient’s glucose level in a normal range (and indeed, blood glucose was frequently not within range, suggesting that the clinician-ordered dose was suboptimal). As designed, our current model attempts to predict the behavior of clinicians and outputs how much insulin the clinician will give to the patients. The predictive capability achievable thus far is not sufficient for a fully automated system but can still provide useful information for clinical decision support.

## Conclusion

Individual blood glucose level and insulin dosing are highly erratic and precise prediction of them is likely impractical. Average blood glucose level over 24 hours can be more reliably predicted and determining whether the patient’s glucose level is going to be high is a more feasible task, which can be sufficient for supporting the discrete decision making in inpatient diabetes management and glucose control.

## Data Availability

This research used data or services provided by STARR, "STAnford medicine Research data Repository", a clinical data warehouse containing live Epic data from Stanford Health Care (SHC), the University Healthcare Alliance (UHA) and Packard Children’s Health Alliance (PCHA) clinics and other auxiliary data from Hospital applications such as radiology PACS. The STARR platform is developed and operated by Stanford Medicine Research IT team and is made possible by Stanford School of Medicine Research Office. The study was approved by the Stanford Institutional Review Board. The content is solely the responsibility of the authors and does not necessarily represent the official views of the NIH, VA, or Stanford Healthcare.

## Acknowledgements

This research was supported in part by the NIH Big Data 2 Knowledge initiative via the National Institute of Environmental Health Sciences under Award Number K01ES026837, the Gordon and Betty Moore Foundation through Grant GBMF8040, and a Stanford Human-Centered Artificial Intelligence Seed Grant. Additional support comes from Diabetes, Endocrinology and Metabolism Training Grant 5T32DK007217-44 and the Enlight Foundation Graduate Fellowship. Patient data were extracted and de-identified by Stanford Medicine’s Research IT department as part of the Stanford Medicine Research Data Repository (STARR) project with support from the Stanford Clinical and Translational Science Award (CTSA) to Spectrum (UL1 TR001085), as led by the National Center for Advancing Translational Sciences at the National Institutes of Health. Additional support is from the Stanford NIH/National Center for Research Resources CTSA award number UL1 RR025744. The content is solely the responsibility of the authors and does not necessarily represent the official views of the NIH or Stanford Healthcare.

